# The successful use of volunteers to enhance NHS Test and Trace contact tracing of in-patients with Covid-19: a Pilot Study

**DOI:** 10.1101/2021.01.28.21250096

**Authors:** Rachel Foster, Bing Jones, Ian Carey, Andzelika Duda, Abigail Reynolds, Jack Czauderna, Alex Westran, Consultant in Infectious Diseases Sheffield Teaching Hospital Foundation Trust

**Author notes:** Corresponding Author: Dr Rachel Foster, Dept Infectious Diseases, Royal Hallamshire Hospital, Glossop Road, Sheffield, S10 2JF, UK.

## Abstract

Contact tracing in the UK for Covid-19 is performed by NHS Test and Trace (NHSTT) via telephone or email. This study estimates how many patients who have been admitted to hospital are not reached by NHSTT and the number of their contacts who were not advised to self-isolate.

Medical Student volunteers conducted face to face interviews with patients diagnosed with Covid-19 on an infectious diseases ward. Data on their close contacts were sent to NHSTT.

20 cases were enrolled. 13(65%) did not engage with NHSTT, 4(20%) because they had no positive PCR, 9(45%) because of severity of illness, language or intellectual difficulties. 49 close contacts were identified of whom 33(67%) were from cases who had not engaged with NHSTT. “Backwards” contacts tracing information was collected from 11(55%) cases and 8(40%) gave detailed information.

These data suggest that NHSTT fails to engage nearly two thirds of Covid-19 in-patients and fails to advise two thirds of their close contacts to self isolate.Volunteers used face to face interviews to overcome false negative tests, illness and communication problems to identify both close contacts and data on sources of infection.

## Introduction

Contact tracing is a fundamental part of controlling the Covid-19 pandemic. Finding the contacts of patients who are in hospital presents a particular challenge to NHS Test and Trace (NHSTT). Senior hospital staff have been aware that many inpatients are too ill to answer phone calls or engage effectively with remote contact tracers. This study aims to estimate how many Close Contacts may be missed. Face-to-face conversations from a prior pilot of 36 inpatients on the ward showed that it was practical to collect details of Close Contacts from Covid-19 patients. Conversations with patients enabled “backward” tracing information on likely sources and locations of infection (including a carvery and social club).

Patients with a clinical diagnosis of Covid-19 but with a negative Polymerase Chain Reaction (PCR) test are not approached by NHSTT, potentially contributing to the spread of infection. There is clearly an unmet need for both backward and forward contact tracing of in-patients. There is provision for contact tracing for hospital-acquired infections but there are no designated resources for contact tracing of those admitted with Covid-19, other than through NHSTT.

## Method

A standard operating procedure was prepared, including a simple pro-forma for data collection. Third year medical student volunteers were recruited as part of their community placements with a local community contact tracing group.^1^ Training included Contact Tracing, use of patient database and ward-based personal protective equipment and procedures. 20 inpatients were identified on infectious diseases wards of a major teaching hospital. Entry criteria were either clinical or laboratory confirmed Covid-19. Clinically diagnosed patients met criteria which included epidemiological, radiological and/or haematological changes consistent with Covid-19. Face to face interviews were conducted on the ward. Data collected included identifiers, testing data, contact with NHSTT, details of close contacts and potential sources of infection. Each proforma was copied and sent to the local health protection team to enable entry onto NHSTT’s Contact Tracing and Advisory

Group (CTAS)^2^ system (see discussion). Data were anonymized and entered onto a spreadsheet for analysis. Patients specifically identified as part of hospital outbreaks were excluded since they are managed separately. One patient is included after testing positive in the community following an outbreak in another hospital.

## Results

20 cases were enrolled. 4 (20%) were awaiting results or PCR negative. 11 (55%) patients had been contacted by NHSTT of whom 7 (35%) engaged and gave details of their close contacts. 13 (65%) did not engage with NHSTT of whom 4 (20%) were either too ill or had language difficulties. A further two (10%) were unsure whether they had been contacted by NHS test and trace because of language or cognitive difficulties. 49 close contacts of all cases were identified of whom 33 (67%) were contacts of cases who had not engaged with NHSTT. “Backwards” tracing information on possible sources of infection was given by 11 (55%) of whom 8 (40%) gave detailed tracking information including individuals, named schools or health and care settings. The time taken to interview and collect data on all 20 cases was 20 hours including the use of personal protective equipment and access to hospital IT. See table:

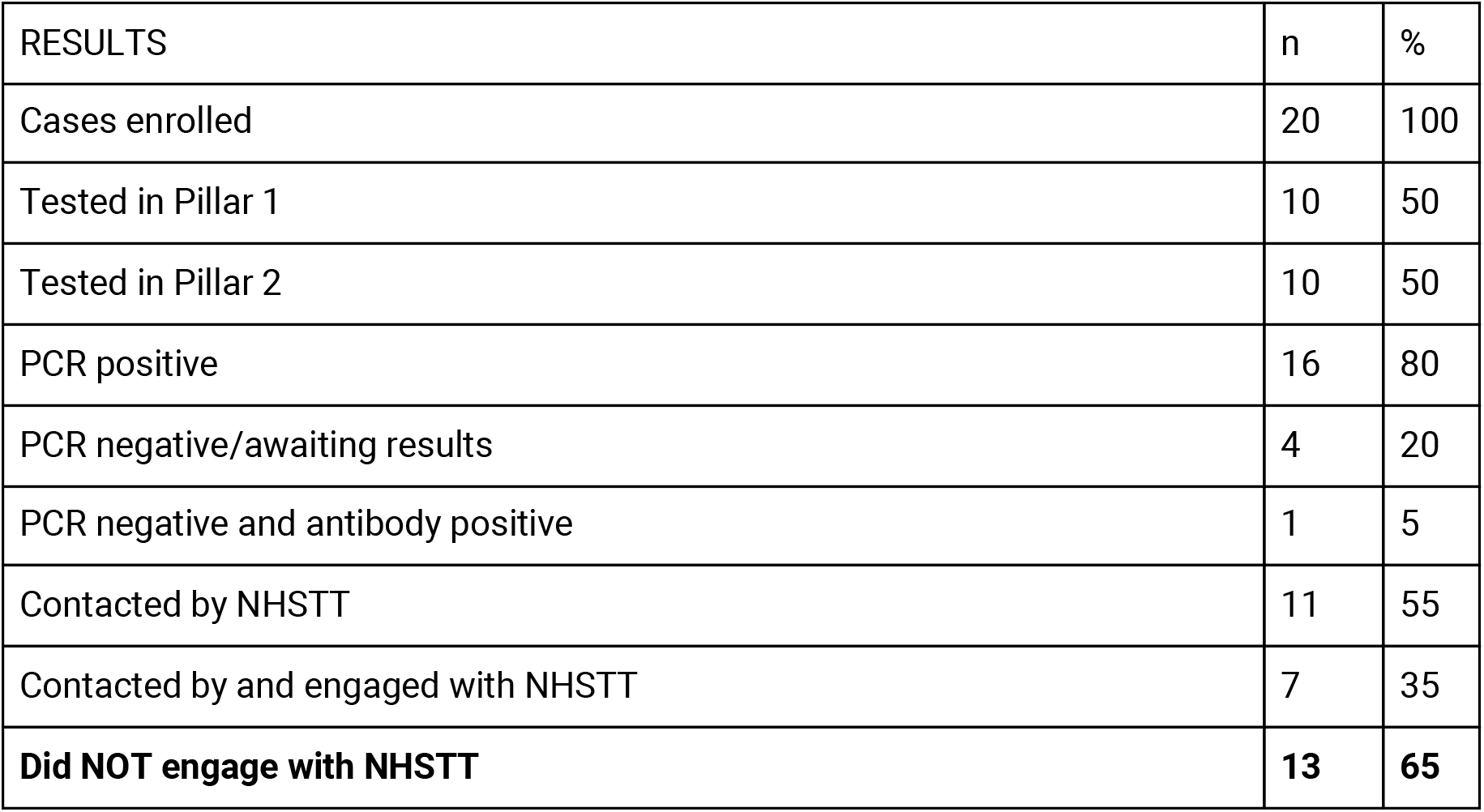
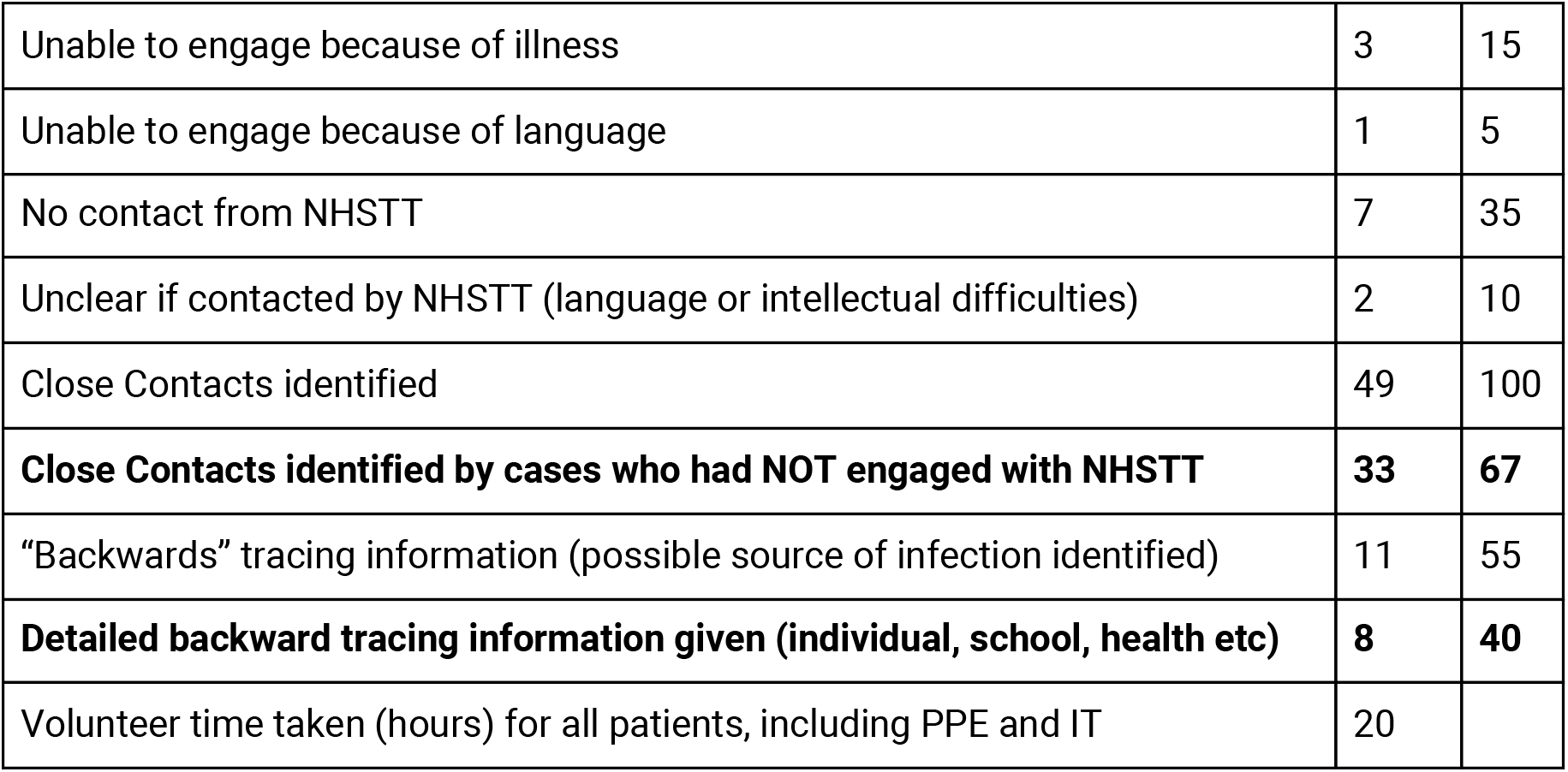

| RESULTS | n | % |
| --- | --- | --- |
| Cases enrolled | 20 | 100 |
| Tested in Pillar 1 | 10 | 50 |
| Tested in Pillar 2 | 10 | 50 |
| PCR positive | 16 | 80 |
| PCR negative/awaiting results | 4 | 20 |
| PCR negative and antibody positive | 1 | 5 |
| Contacted by NHSTT | 11 | 55 |
| Contacted by and engaged with NHSTT | 7 | 35 |
| **Did NOT engage with NHSTT** | **13** | **65** |
| Unable to engage because of illness | 3 | 15 |
| Unable to engage because of language | 1 | 5 |
| No contact from NHSTT | 7 | 35 |
| Unclear if contacted by NHSTT (language or intellectual difficulties) | 2 | 10 |
| Close Contacts identified | 49 | 100 |
| **Close Contacts identified by cases who had NOT engaged with NHSTT** | **33** | **67** |
| "Backwards" tracing information (possible source of infection identified) | 11 | 55 |
| **Detailed backward tracing information given (individual, school, health etc)** | **8** | **40** |
| Volunteer time taken (hours) for all patients, including PPE and IT | 20 |  |

## Discussion

Mortality from Covid-19 has been exceptionally high in the UK^3,4,5^. Lockdowns, mass testing and vaccination strategies are important control strategies but it is likely that the UK will remain in need of effective contact tracing for many months. NHSTT is flawed, missing the untested, false negatives and those cases it fails to reach. Many of the cases missed by NHSTT will be admitted to hospital. More than 80% of these potentially infectious contacts may be missed at present ^6^. Contact tracing by NHSTT can be delayed^7^. Inadequate or delayed contact tracing of Covid-19 inpatients results in avoidable ongoing transmission.

Contact tracing of in-patients is a particular problem which has received little national publicity. The report authors have not identified any similar work being undertaken in the UK. During August the total number of Covid-19 inpatients was less than 1,000, but had risen to circa 14,000 during the week of data collection in December 2020^8^.

This study demonstrated that **65% of the inpatients studied had not engaged with NHSTT**. Particular contact tracing problems for in-patients were identified, with 35% of patients failing to engage with NHSTT because of illness, language, hearing or cognitive barriers. Face to face inpatient contact tracing overcomes these barriers. This study also engaged 20% of cases with a clinical diagnosis but no laboratory confirmation, enabling immediate collection of tracing data. Failure to detect untested and false negative community cases are also limitations of NHSTT, but are more difficult to resolve.

33 close contacts were identified from the 13 (65%) of cases that NHSTT had failed to engage. This suggests that **67% of all the contacts identified were not traced by NHSTT**. If extrapolated, this suggests a major omission nationally. The Contacts of patients ill enough to be admitted may be more likely to effectively self-isolate than the disappointing current estimate of 2 0%^9^.

This study found 20% of cases with a clear clinical diagnosis were awaiting results or had tested PCR negative. Currently, these cannot be detected by NHSTT or entered onto the national computerised (CTAS)^10^ system. This is despite a current legal requirement for suspected cases to be notified^11^ to the Proper Officer.

Solutions need to be explored to enable clinically diagnosed and false negative patients to be registered to enable the tracing of their contacts.

Data on contacts were transferred to the City Council Contact Tracing service who had difficulties using information because they can only add data if patients already have an NHSTT computerised record. This means that inpatients and their contacts that have been missed by NHSTT cannot be registered by NHSTT. Local authority staff have repeatedly complained about this feature of CTAS, the national IT system. Financial and other support, normally available, cannot be offered to these index cases or their contacts.

This study demonstrates that medical student volunteers can effectively conduct face-to-face interviews in a hospital setting. Volunteers reported patients and their relatives were willing to engage for the wider good. Patients expressed gratitude for the time spent listening to their stories. Volunteers effectively collected email or phone numbers for a majority of Close Contacts. The identity of these Contacts were successfully referred to NHSTT, through the City Council Public Health Team for entry into CTAS and subsequent contact tracing of those with a positive PCR by NHSTT.

This study demonstrates that face to face interviews can overcome particular problems relating to contact tracing of inpatients by being sensitive to symptoms, circumstance, and by employing non-verbal communication (including signing/pointing) and translation software.

There is increasing evidence that backward contact tracing is effective^12^. In a preliminary exercise, one of the authors identified a particular public house as a common source of infection, enabling intervention by the local environmental health team. In this study, 55% of cases offered information on possible sources of infection. Three patients had been exposed to positive household members, one had been in a care home outbreak. One had been in an outbreak in another hospital, detected in the community after discharge and not contacted by NHSTT. If coordinated with local knowledge these data could be crucial for outbreak control.

This was a small proof of principle study undertaken in a short time frame with zero budget. Medical student volunteers did not undertake contact tracing themselves but if this could be coordinated with other public health services it could be a timely and effective asset. The use of volunteers may not be sustainable and salaried personnel should be considered.

This proof of principle study demonstrates the urgent need for hospital based inpatient contact tracing. It suggests that a significant number of cases and contacts that are currently missed by NHSTT could be advised to self-isolate. It suggests that a volunteer-based service is practical and effective. It demonstrates a need for increased flexibility and connectivity with local Health Protection Teams and NHSTT.

## Data Availability

All data available from the corresponding author

## Competing interests

All authors have completed the ICMJE uniform disclosure form at www.icmje.org/coi_disclosure.pdf and declare: no support from any organization for the submitted work, no financial relationships with any organizations that might have an interest in the submitted work in the previous three years, no other relationships or activities that could appear to have influenced the submitted work.

## Authors

Rachel Foster, Consultant in Infectious Diseases Sheffield Teaching Hospital Foundation Trust (Correspondence to)

Ian Carey, RGN, Sheffield, working as a volunteer

Andzelika Duda, 3rd year medical student University of Sheffield

Abigail Reynolds, 3rd year medical student University of Sheffield

Jack Czauderna, retired GP Sheffield

Bing Jones, retired Associate Specialist in Haematology Sheffield

Alex Westran, Operational Manager for Sheffield City Council Track and Trace.

References are hyperlinked and in footnotes.

## Footnotes

1 Sheffield Community Contact Tracers [online] Available at: http://www.communitycontacttracers.com/ [accessed 02/01/2021]

2 Gleave, R. (24th April 2020). *Letter to DsPH on contact tracing*. London: Public Health England

3 European Centre for Disease Prevention and Control, *COVID-19 situation update worldwide, as of week 50 2020*. Available from: https://www.ecdc.europa.eu/en/geographical-distribution-2019-ncov-cases [accessed 21st December 2020]

4 Public Health England, *Coronavirus in the UK // Deaths*. Available from: https://coronavirus.data.gov.uk/details/death [accessed 21st December 2020]

5 Public Health England, *Coronavirus in the UK // Cases*. Available from: https://coronavirus.data.gov.uk/details/cases [accessed 21st December 2020]

6 Jones, B; Czauderna, J; Regrave, P. (2020) *We must stop being polite about Test and Trace—there comes a point where it becomes culpable*. London: BMJ Publishing Group Limited 2020.

7 Davies, G. (11th December 2020). *The government’s approach to test and trace in England - interim report*, p13. National Audit Office.

8 NHS (2020). *COVID-19 Hospital Activity*. [online] Available at: https://www.england.nhs.uk/statistics/statistical-work-areas/covid-19-hospital-activity/ [accessed 23/12/2020]

9 Smith, L et al. (2020) *Adherence to the test, trace and isolate system: results from a time series of 21 nationally representative surveys in the UK (the COVID-19 Rapid Survey of Adherence to Interventions and Responses [CORSAIR] study)*. medRxiv [online]. Available at: https://www.medrxiv.org/content/10.1101/2020.09.15.20191957v1 [accessed 23/12/2020]

10 Gleave, R. (24th April 2020). Letter to DsPH on contact tracing. London: Public Health England

11 Department of Health and Social Care (5th March 2020). Coronavirus (COVID-19) listed as a notifiable disease. [online] Available from: https://www.gov.uk/government/news/coronavirus-covid-19-listed-as-a-notifiable-disease [accessed 23/12/2020]

12 Endo A et al. (October 2020) *Implication of backward contact tracing in the presence of overdispersed transmission in COVID-19 outbreaks*. Wellcome Open Res 2020, 5:239 (https://doi.org/10.12688/wellcomeopenres.16344.1)

## Notes

### Competing Interest Statement

The authors have declared no competing interest.

### Clinical Trial

Small scale ward based pilot study.

### Funding Statement

This was an unfunded study.

### Author Declarations

A waiver of ethical approval was received from Research and Innovation department at Sheffield Teaching Hospitals NHS Foundation Trust.

